# GHSI COVID-19 puzzle: did highly developed countries indeed fare worse?

**DOI:** 10.1101/2022.08.28.22279258

**Authors:** Sofija Markovic, Igor Salom, Andjela Rodic, Marko Djordjevic

## Abstract

Global Health Security Index (GHSI) categories are formulated to assess the capacity of world countries to deal with infectious disease risks. Thus, higher values of these indices were expected to translate to lower COVID-19 severity. However, it turned out to be the opposite, surprisingly suggesting that higher estimated country preparedness to epidemics may lead to higher disease mortality. To address this puzzle, we: *i*) use a model-derived measure of COVID-19 severity; *ii*) employ a range of statistical learning approaches, including non-parametric machine learning methods; *iii*) consider the overall excess mortality, in addition to official COVID-19 fatality counts. Our results suggest that the puzzle is, to a large extent, an artifact of oversimplified data analysis and a consequence of misclassified COVID-19 deaths, combined with the higher median age of the population and earlier epidemics onset in countries with high GHSI scores.

## Introduction

It is natural to assume that the most prosperous world countries, well organized and with the most advanced medical institutions, would soon gain an advantage in the global race to minimize the harm from a new viral infection. This expectation is quantified in the form of the Global Health Security Index (GHSI) - a measure of preparedness for various health emergencies, precisely of the kind of the present COVID-19 pandemic ^1^. The categories included in this index were carefully designed to help countries identify weaknesses primarily in their healthcare systems but also in their political and socio-economic structure, as well as to guide the process of reinforcing their health security capacities ^2–4^. To this end, capacities to prevent medical hazards, to timely detect and report them, to rapidly respond in these situations, to comply in all these with international norms, as well as the health system capacity, and the overall vulnerability to biological threats - are all separately assessed and assigned a specific GHSI category: Prevent, Detect, Respond, Norms, Health and Risk, respectively.

With the COVID-19 pandemic, the GHSI scores were put for the first time to a real-life global test – which they failed, according to most of the published attempts to assess their correlation with the epidemic scale ^5–18^. An unexpected positive correlation between the GHSI categories and differently calculated mortality rates ^5–7,12,14,17^ and infection spread ^5,7,8,10,12–14^ – deserving to be called the “GHSI puzzle” – suggested that the GHSI cannot be used to describe the national pandemic preparedness ^19^. As this result seemed highly convincing, the focus was shifted to the search for explanations of the GHSI inappropriateness ^19^. Thereby, the observed large COVID-19 infection counts in highly developed (high GHSI countries) motivated the firm opinion that their responses were “sclerotic and delayed at best” ^20^. The first-ranked country in the GHSI 2019 list, the USA, provided a striking example. Despite its top overall score of 76/100 - equaling roughly twice the average score of 38.9 - COVID-19 has taken a devastating toll in the US, especially in the first wave of the pandemic ^21,22^, introducing a two-year drop in life expectancy ^23^. It was pointed out that GHSI scores cannot anticipate how a country decides to respond to epidemics, so these scores cannot be expected to necessarily be predictive of reported infections and fatalities ^24,25^.

However, it is also necessary to revisit the methodological approaches that lead to the unintuitive relation of GHSI scores to the observed pandemic effects. First, the outcome variable, assumed to depend on the GHSI, should measure the relevant, effective epidemic burden to reasonably represent the country’s overall success in minimizing the harm from the epidemic. Significantly, it should predominantly depend on the aspects of preparedness rather than other prevailing factors. As noted above, the GHSI predictivity was analyzed for different mortality and infection rates, yielding mostly positive correlations, and these variables depend on factors that directly or indirectly increase the frequency and range of personal contacts ^26^ and thus promote the disease spread ^27^. Developed countries, typically carrying high GHSI scores, are characterized by intense population mobility and mixing due to ease of travel, higher local concentrations of people, and business and social activities. So, the consequential rapid disease spread may cancel and overshadow the ameliorating effects of large preparedness capacities, explaining the lack of a negative correlation of the GHSI with COVID-19 growth numbers. In addition, these outcome variables depend on the calculation time frame, and there are even examples of incidence rates with negative dependence on the GHSI ^18^. Therefore, it is quite nontrivial to select the appropriate outcome variable (among mortality and incidence rates) for describing the relationship between the GHSI and the epidemic burden.

The relation of the GHSI with clinical severity measured by the Case Fatality Ratio (CFR), as a more obvious indicator of the burden magnitude, was considered in a few papers ^8,13,16^, with the most extensive study (for a large number of countries and with including numerous other potential predictors) carried out in ^13^ - and the connection turned out to be positive. This is genuinely perplexing since CFR is an objective measure of disease severity, thus expected to be causally independent of the disease transmissibility. Consequently, there is no obvious explanation why higher GHSI would be linked to a worse case fatality ratio.

While CFR is a specific measure of severity, its shortcoming is being time-dependent and affected by the varying lag between case detections and fatal outcomes (in addition to lacking a direct mechanistic interpretation in terms of the disease dynamics) ^28^. Instead, we choose a timing-independent measure of severity: m/r value that we previously derived from an epidemic model ^29^, which represents the ratio of population-averaged mortality to recovery rates (so that many fatalities and/or slow recovery correspond to higher epidemic severity). It can be shown that this measure is independent of the SARS-CoV-2 transmissibility and directly relates to the asymptotic CFR value calculated at the end of the epidemic wave, so it does not rely on an arbitrary time frame. Thus, we could estimate it straightforwardly using the data from the end of the first pandemic’s peak, avoiding interpretation of complex effects of vaccination and different SARS-CoV-2 strains on the disease severity.

Furthermore, we aimed to check if the observed positive GHSI correlations with various measures of epidemics intensity may be an artifact of the applied methods that are not well adapted for the problem complexity – i.e., cannot extract the impacts specific to individual factors entangled in mutual correlations. To that end, we searched for the m/r predictors among a number of variables on 85 countries using methods such as the regularization-based linear regressions (Lasso and Elastic Net) ^30^, and non-parametric machine learning techniques (Random Forest and Gradient Boost) ^30^. Together with different GHS Indices, we also included a number of sociodemographic and health-related variables in the analysis, in line with a recent result that factors external to GHSI may aid in characterizing countries’ ability to respond to epidemic outbursts ^31^. Finally, we investigated the relationship between the GHSI and the total excess deaths, which was not performed before; the main advantage of such analysis is that, unlike official COVID-19 figures, total excess deaths counts do not depend on diagnosis and reporting policies ^17,18^. Combined together, our results, to a large extent, solve - or more precisely dissolve - the puzzle: it is not justified to relate high GHSI values (i.e., high estimated country preparedness) with an unsatisfactory pandemic response and high severity/mortality burden.

## Methods

The data was acquired from several online databases (for details, see Supplementary Material). Briefly, Global Health Security Indices data were taken from ^1^, total and excess deaths data during the first peak of the pandemic were obtained from ^32^, while COVID-19 counts and demographic data were collected similarly to those previously used in ^27^. A measure of disease severity – mortality over recovery rate (m/r) – was inferred for each country, following the methodology introduced in ^29^. Relative excess deaths and relative unexplained deaths (excess and unexplained deaths in further text and figures) were calculated as follows:

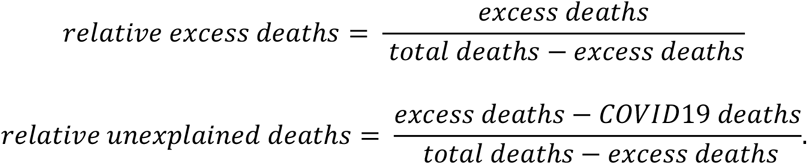

Data processing and subsequent univariate and multivariate statistical analysis, including regularization-based linear regressions (Lasso and Elastic Net) ^30^, and non-linear machine learning techniques (Random Forest and Gradient Boost) ^30^, employed similar methodology previously used in ^27,29^ - see a detailed description in Supplementary Material. The contribution of the relevant variables in Random Forest and Gradient Boost was estimated through Partial Dependence (PD) plots ^30^, which estimate the effect of a single predictor by taking into account the average effects of all other predictors.

Supervised Principle Component Analysis (PCA) ^30^ was performed. In this method, PCA is done only on those variables that show sufficiently high correlation with the response variable (m/r), where a certain number of PCs is then used in multiple regression with m/r as the response. The univariate correlation constant cutoff (i.e., the number of variables retained for PCA), and the number of retained PCs, are treated as hyperparameters, i.e., determined through cross-validation. Principal components that appeared significant in the multivariable regressions were further analyzed.

Hyperparameter optimization was performed for all machine learning methods through five-fold cross-validation (data was repartitioned 40 times). A combination of hyperparameters, leading to the minimum mean squared error on the testing set, was used in the final model. These selected hyperparameter values were then used to train the models on the entire dataset.

## Results

### Analysis based on COVID-19 case counts

Univariate analysis (Fig. 1) reveals a statistically significant positive correlation of all selected GHSI categories with our severity measure (m/r), demonstrating the puzzle. Strikingly, GHSI categories show the highest positive correlations compared to all other variables in the analysis. Also, they are highly correlated with measures of a country’s development (e.g., GDP per capita and HDI), so that, on average, more developed countries have higher GHSI.

**Figure 1:**
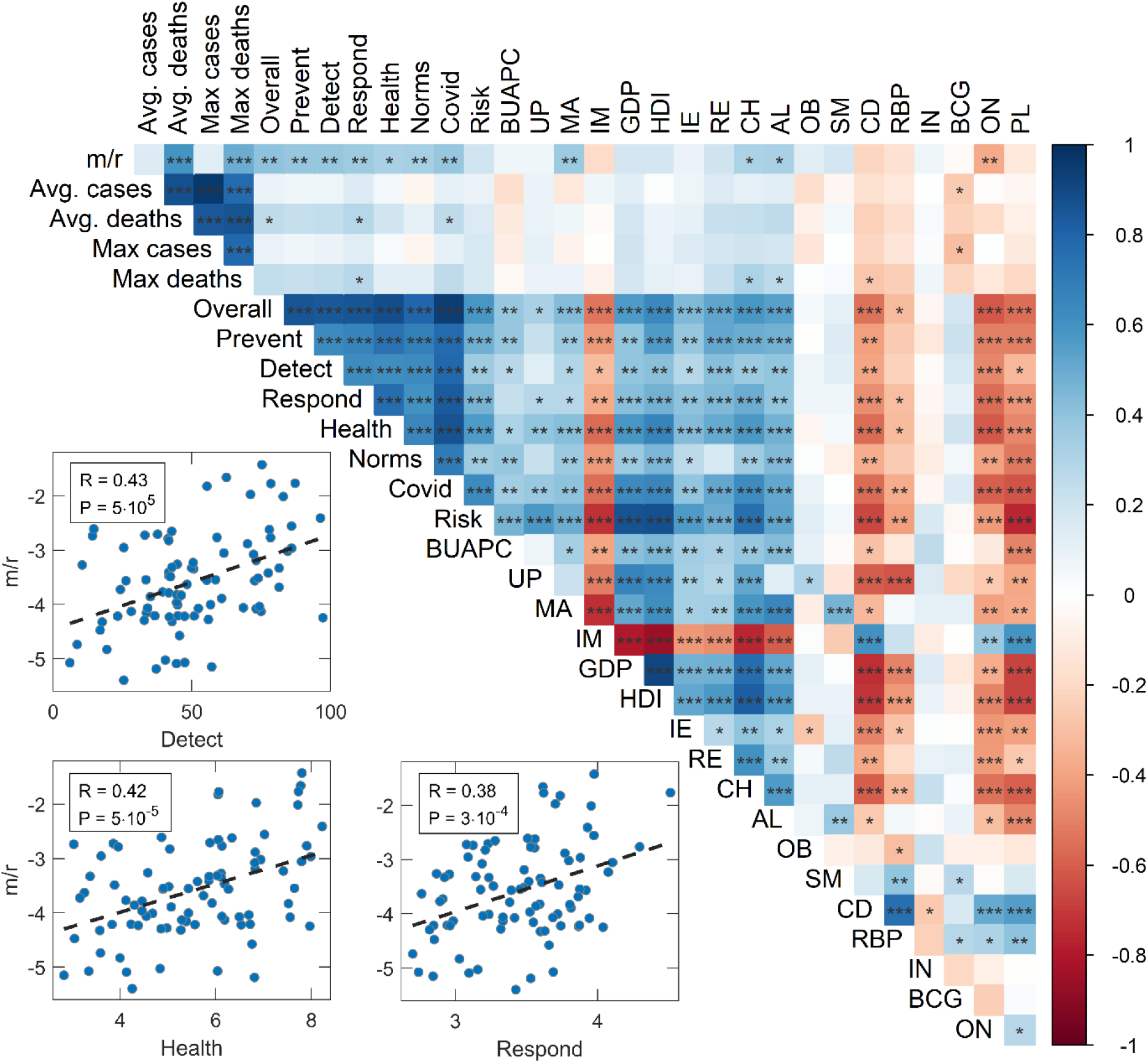
Graphical representation of the predictor correlation matrix. (as the matrix is symmetric, only its upper half is shown). Colors correspond to the magnitude of Person correlation coefficients, as indicated by the color bar on the right, while asterisks correspond to the statistical significance of the results (‘ ‘ – P>0.05, ‘ * ‘ – 0.05>P>0.01, ‘ ** ‘ – 0.01>P>0.001, ‘ *** ‘ – P< 0.001). Correlations of m/r and selected three Global Health Security Index categories, relevant for the pandemic, are also represented by scatterplots in the inset. m/r – disease severity measure, Overall – Overall GHSI, Prevent – GHSI Prevent category, Detect – GHSI Detect category, Respond – GHSI Respond category, Health – GHSI Health category, Norms – GHSI Norms category, Risk – GHSI Risk category, Covid – a combination of COVID-related GHSI indicators, BUAPC – built-up area per capita, UP – urban population, MA – median age, IM – infant mortality, GDP – gross domestic product per capita, HDI – human development index, IE – net migration, RE – refugees, CH – blood cholesterol level, AL – alcohol consumption, OB – prevalence of obesity, SM – prevalence of smoking, CD – prevalence of cardiovascular diseases, RBP – raised blood pressure, IN – physical inactivity, BCG – BCG vaccination coverage, ON – the onset of the epidemic, PL – air pollution. The figure was created using R ^33^ corrplot package ^34^, in RStudio integrated development environment for R ^35^.

However, the univariate analysis does not control for simultaneous effects of multiple factors that can influence the disease severity. Additionally, many factors that we initially included as potentially important (and thus considered in the analysis) may not significantly influence the severity, so retaining them in the final model would introduce large noise. Another problem is that many factors are mutually highly correlated. Consequently, we next applied linear regressions with regularizations and feature selection (Lasso and Elastic Net, which can eliminate non-significant predictors), combined with PCA on groups of related variables (to partially decorrelate the variable set while retaining its interpretability).

Mutually related variables were thus grouped in three categories (age, chronic disease, prosperity) on which PCA was performed (Supplement Fig 1, Supplement Table 2), while the other variables remained unchanged (Supplement Table 1). The data were then independently analyzed using Lasso (Fig. 2A) and Elastic net regressions (Fig. 2B), which were implemented in the so-called “relaxed” ^30^ way to reduce noise from the multidimensional data.

**Figure 2:**
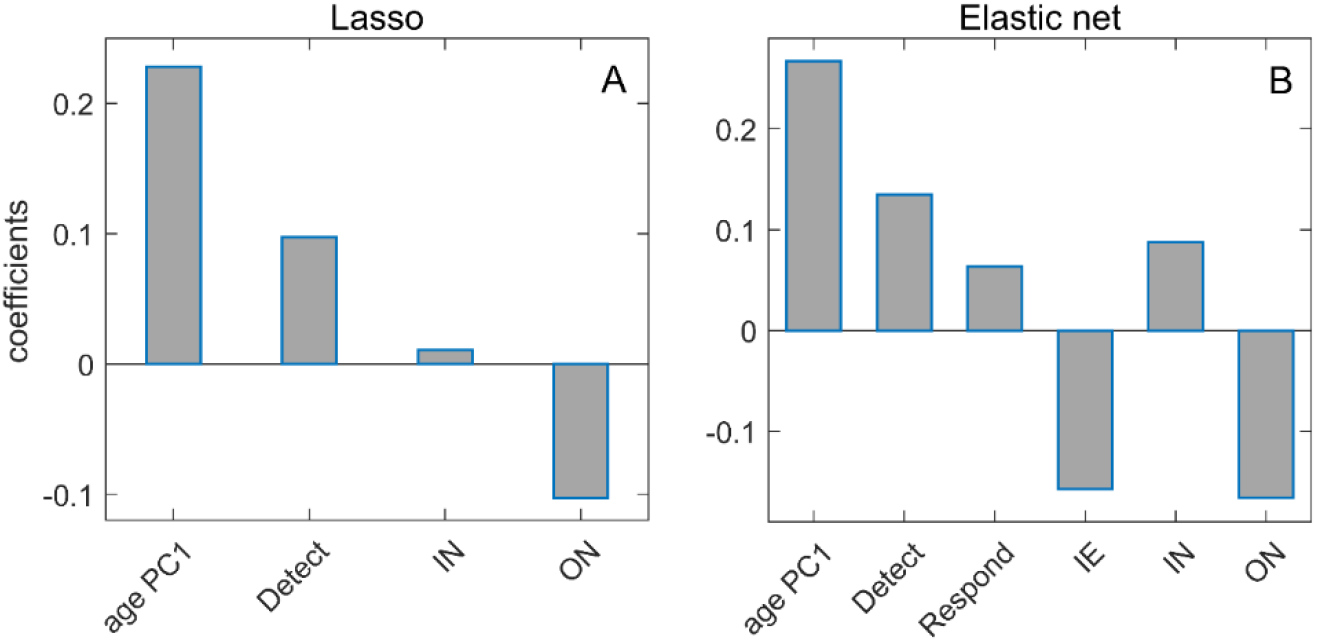
Lasso and Elastic Net regressions with m/r as the response variable. A) Regression coefficients of variables selected by Relaxed Lasso regression, B) Regression coefficients of variables selected by Relaxed Elastic Net regression, age PC1 – age principal component 1, Detect – GHSI Detect category, Respond – GHSI Respond category, IE – net immigration, IN – physical inactivity, ON – the onset of the epidemic.

From Fig. 2, we see that age PC1 is the dominant predictor in both methods, unsurprisingly suggesting that the older population is more severely affected by COVID-19. Furthermore, epidemic onset appears to be an important factor negatively associated with the disease severity, meaning that earlier epidemic onset is associated with higher severity of the disease. Of all four analyzed GHSI categories, the disease detection index is selected as the most important, with an (unintuitive) positive effect on m/r. While positive contribution was also obtained through univariate analysis, its magnitude diminishes when the more advanced analysis is used, i.e., in Fig. 2, PCA followed by multivariate regressions with regularization. Consequently, while the paradox is still there (positive contribution of one of GHSI categories), the regression coefficient of this predictor significantly decreases (e.g., compared to age) as the analysis progresses from univariate to multivariate (with feature selection and correlated variables accounted for). Also, in Elastic net regression (Fig. 2B), net immigration appears to contribute negatively to the disease severity, which we will further discuss below. The obtained results remain robust when the regression methods are applied to the initial data without PCA, when besides 18 demographic parameters, Detect, Respond, Health and Risk GHSI categories (Supplement Fig. 2), or Covid index (Supplement Fig. 3) are used.

**Figure 3:**
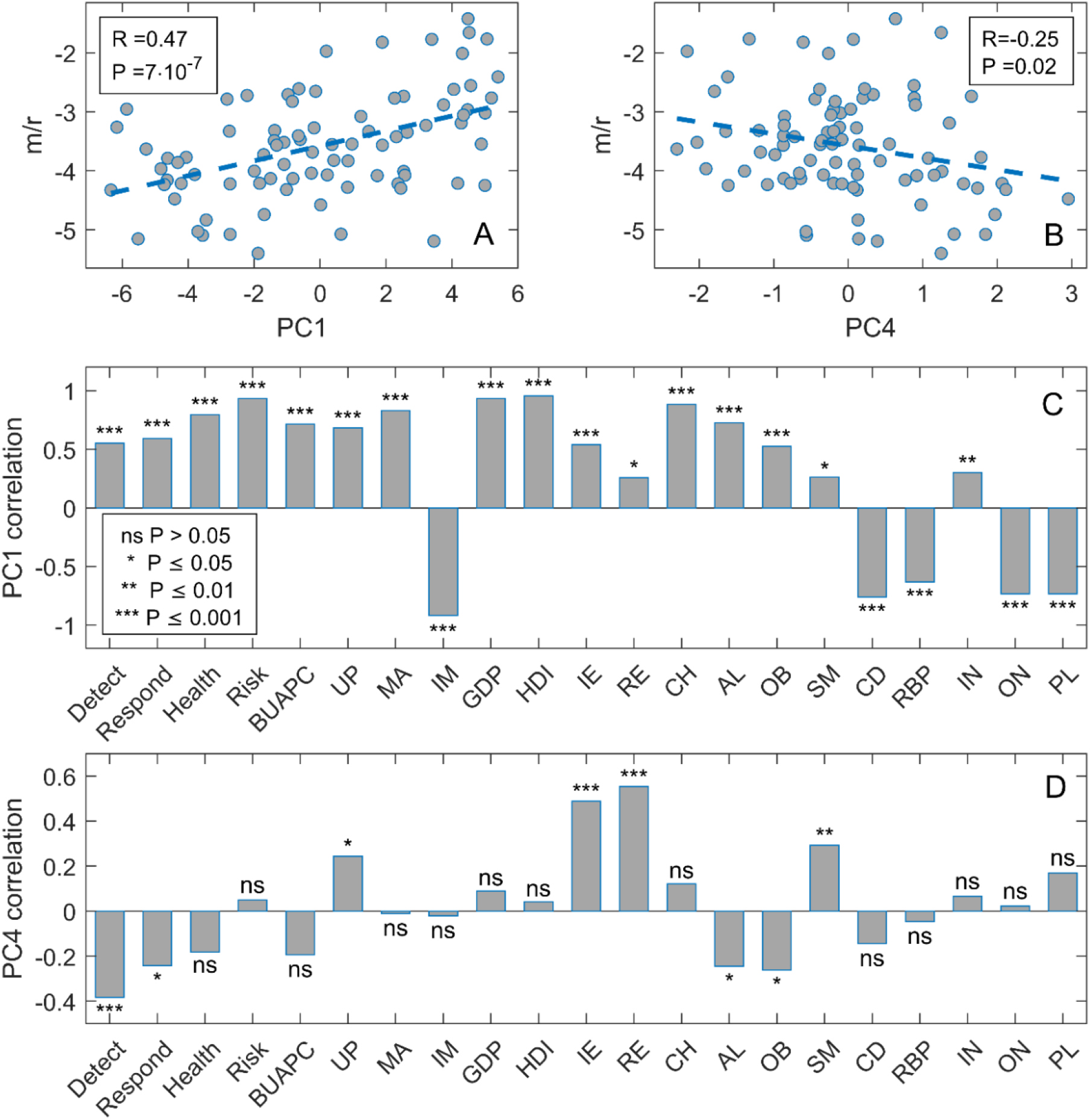
Supervised Principal Component Analysis. Scatterplot of A) PC1 and m/r, B) PC4 and m/r, C) PC1 and variables entering PCA, D) PC4, and variables entering PCA. m/r – mortality over recovery rate, Detect – GHS Detect category, Respond – GHSI Respond category, Health – GHSI Health category, Risk – GHSI Risk category, BUAPC – built-up area per capita, UP – urban population, MA – median age, IM – infant mortality, GDP – gross domestic product per capita, HDI – human development index, IE – net immigration, RE – refugees, CH – blood cholesterol level, AL – alcohol consumption, OB – prevalence of obesity, SM – prevalence of smoking, CD – prevalence of cardiovascular diseases, RBP – raised blood pressure, IN – physical inactivity, BCG – BCG vaccination coverage, ON – the onset of the epidemic, PL – air pollution.

In the analysis above, we grouped variables in subsets according to their natural interpretation (e.g., age-related, chronic diseases, country prosperity) for PCA. While this approach facilitates the interpretation of the regression results, there is also some arbitrariness in the grouping criteria. This motivated us to perform supervised PCA - a mathematically well-defined procedure that chooses the variables for PCA solely based on their numerical correlation with the response variable. The results in Fig. 3 show that two significant principal components remain in the linear regression model – PC1 and PC4. One can see a high correlation of m/r with PC1, which technically makes it a good risk index for COVID-19 severity – though some of its constituents, such as GHSI, have an unintuitive contribution to m/r. More importantly, PC1 shows that the composite variable, which can explain much of the variability in m/r, contains GHSI categories intertwined with HDI and GDP in terms of their effect on m/r. Consequently, the unintuitive positive contribution of GHSI to m/r (as well as the absence of negative contribution in more complex models) may be an indirect consequence of some effect that increases m/r for more developed countries (for which both HDI/GDP and GHSI are high). We will further explore this possibility through the analysis of excess deaths.

The regression methods used so far do not account for nonlinearities or possible interactions between predictor variables. For this reason, we applied machine learning methods based on ensembles of weak learners (decision trees), Random Forest, and Gradient Boost, on the same set of predictors and PCs used in the linear regression models above. Figure 4 shows the estimate of variable importance (A and D) and Partial Dependence (PD) plots (B, C, E, F) for the variables with the highest importance in explaining disease severity. PD plots also provide information about the direction (positive or negative) in which the predictors affect m/r. We obtain age PC1 as by far the most important variable, where older age promotes the disease severity. Detect GHSI and epidemic onset also appear above the standard importance threshold. However, the estimated effect of Detect GHSI is now small, based on both the variable importance and PD plots that show a much larger dependence of m/r on age PC1.

**Figure 4:**
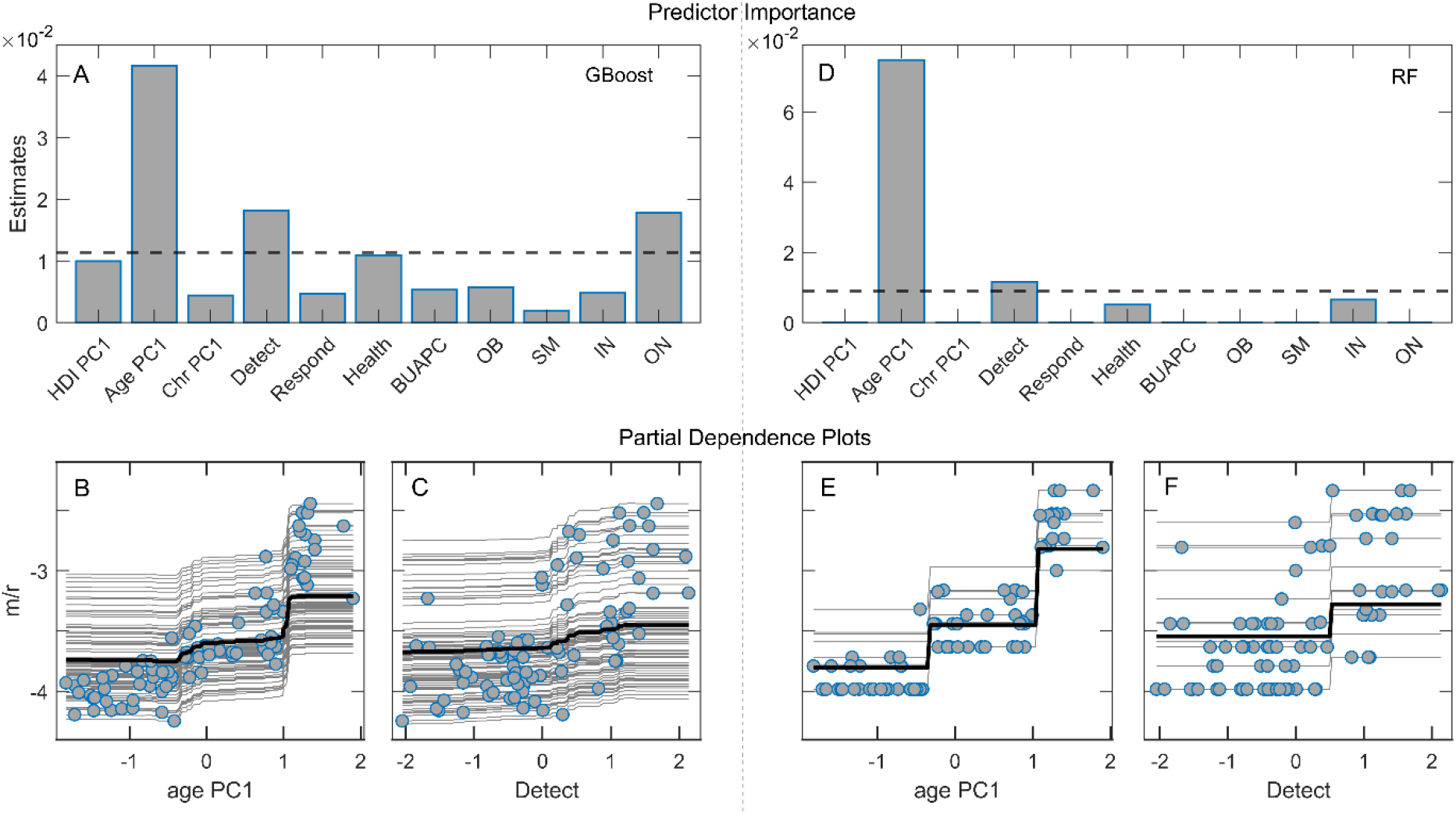
Machine learning algorithms and Partial dependence (PD) plots with disease severity as the response variable. A) Variable importance estimates for Random Forest regression, B-C) PD plots corresponding to Random Forest, D) Variable importance estimates from Gradient Boost regression, E-F) PD plots corresponding to Gradient Boost. **The dashed lines in A and D correspond to the mean variable importance in the given model**. HDI PC1 – human development index PC1, Chr PC1 – chronic diseases PC1, Detect – GHSI Detect category, Respond – GHSI Respond category, Health – GHSI Health category, BUAPC – built-up area per capita, OB – obesity, SM – smoking, IN – physical inactivity, ON – epidemic onset, m/r – disease severity measure.

### Analysis of excess deaths

We next focus on excess deaths (see the definition in Methods), where we again use several univariate and multivariate approaches. As predictors, we here use COVID-19 counts, GHSI, and demographic variables, which we separated into mutually related subsets on which PCA was performed (see Supplement Table 3 and Supplement Fig. 4). In Fig. 5, which shows Random Forest results, we see that Counts PC1, which equally well reflect COVID-19 infection and death case counts, are by far the most dominant predictor of excess deaths. While expected, this is also a highly nontrivial result, given that the two quantities (excess deaths and classified COVID-19 deaths) are inferred in an entirely independent manner, and that in-principle ununiform policies of COVID-19 related deaths classification are applied in different countries ^36^. This result is robust, i.e., consistently obtained through different methods, ranging from simple univariate regressions to linear regressions (with variable selection and regularization), see Supplement Figs. 5 and 6 and Supplement Table 4. The only other predictor with importance above the threshold in Fig. 5 is CFR (which we also calculate in saturation, i.e., at the end of the first wave), which has a small to moderate positive effect on m/r, though much smaller than the COVID-19 case counts.

**Figure 5:**
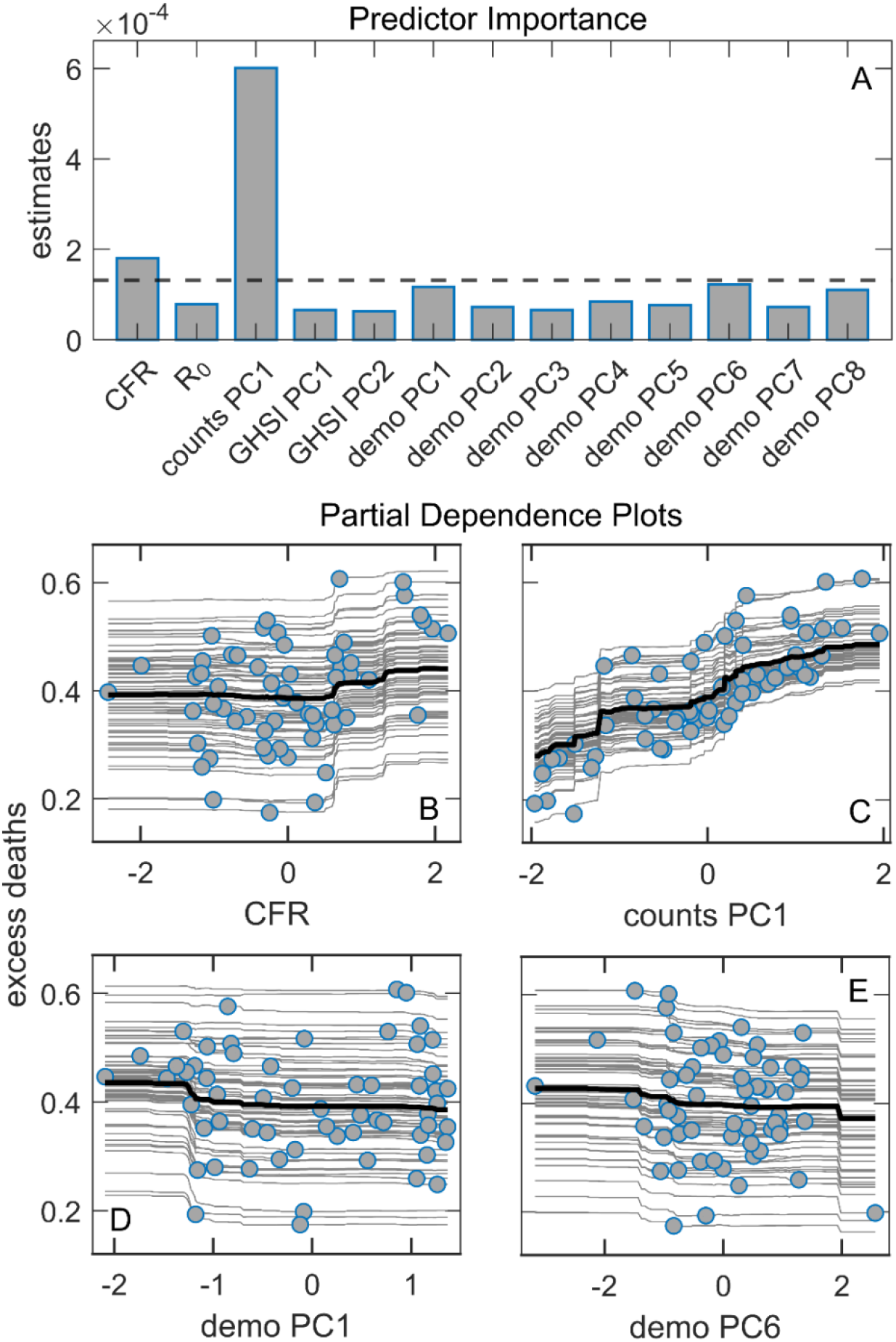
Random Forest regression with excess deaths as the response. A) Predictor importance estimates from Random Forest regression, where dashed line represents the importance threshold, i.e., the mean importance value, B-C) Partial dependence plots of predictors with the highest importance estimates from Random Forest regression model, CFR – case fatality rate, *R*_0_ – basic reproductive number of the virus.

**Figure 6:**
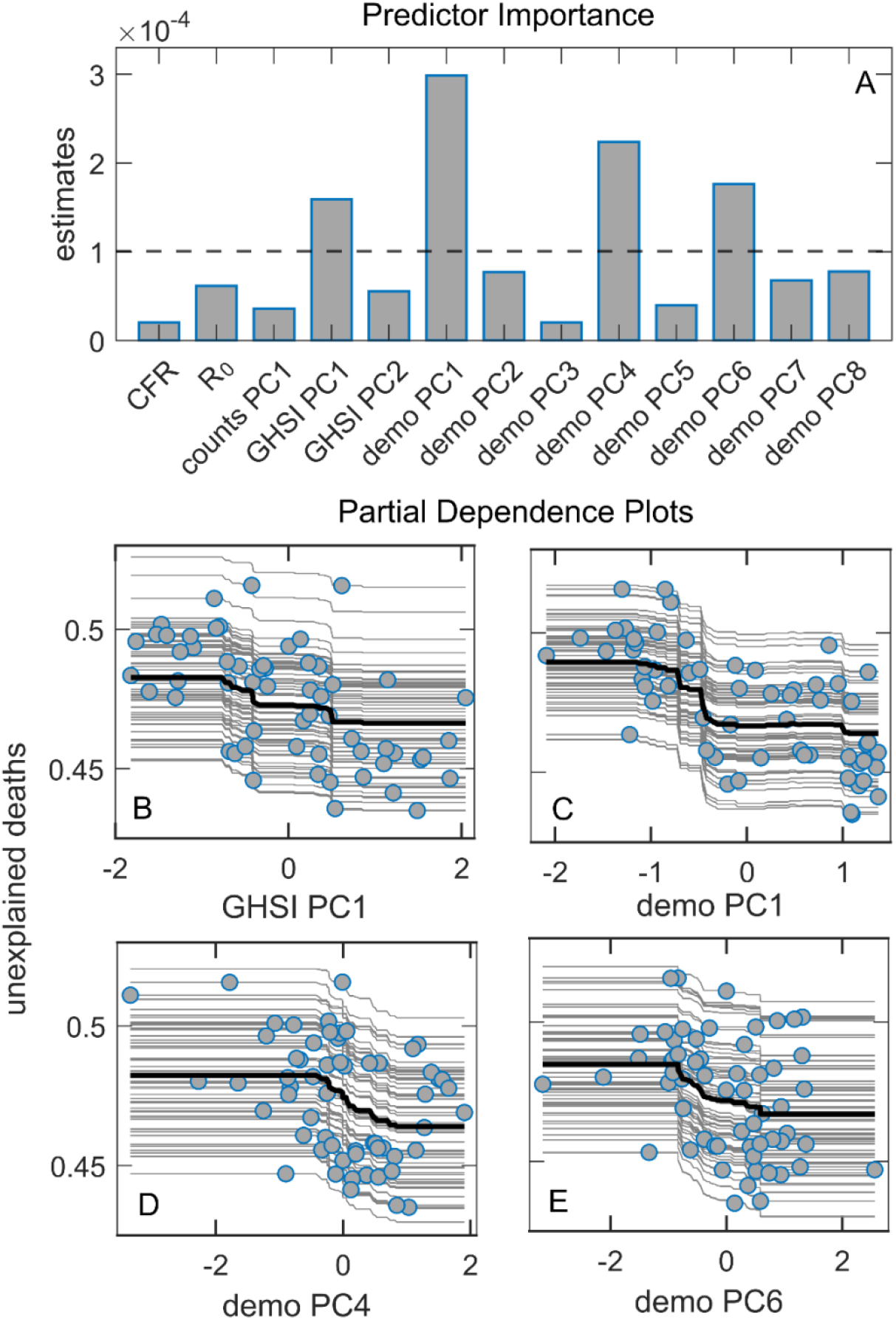
Random Forest regression with unexplained deaths as the response. A) Predictor importance estimates from Random Forest regression, where dashed line represents the importance threshold (the mean value of the variables), B-E) Partial dependence plots od predictor with the highest importance estimates from Random Forest regression model, CFR – case fatality rate, *R*_0_ – basic reproductive number of the virus.

We then analyzed the unexplained deaths with the same approach described above for the excess deaths. The unexplained deaths are the difference between the excess deaths and COVID-19 deaths, i.e., those excess deaths not attributed to COVID-19 (for the definition, see Methods). From Fig. 6, we see that the most important predictor is demo PC1, which has a clear *negative* association with unexplained deaths (see the corresponding PD plot). These results are robust, i.e., independently obtained by several different univariate and multivariate analyses (Supplement Figs. 7 and 8 and Supplement Table 5). As demo PC1 is strongly related to country GDP/HDI (see Supplement Fig. 4), more developed countries are associated with a smaller number of unexplained deaths. The other demographic-related PC components above the importance threshold (demo PC 4 and 6) have a smaller influence on unexplained deaths and can also be related to the negative influence of HDI/GDP (see Supplement Fig. 4). Interestingly, GHSI PC1 (highly correlated to all GHSI categories) is also selected as a significant predictor, negatively associated with unexplained deaths. This implies that higher GHSI (significantly correlated to HDI/GDP) is associated with fewer unexplained COVID-19 deaths.

## Discussion

Although the m/r quantity does not depend on the epidemic growth rate, which is boosted by the large contact rate in countries with high GDP, HDI, and GHSI, the univariate analysis shows that GHSI categories significantly correlate with this measure of severity – having the highest correlation of all considered variables. This could be naively interpreted as a higher likelihood of dying from COVID-19 in well-equipped medical facilities than in settings ranked by far lower GHSI. In the subsequent two parallel analyses - Lasso and Elastic Net on PCs obtained from three predefined groups of variables, on one side, and the Supervised PCA, on the other – the positive effect of GHSI is acknowledged, primarily through the Detect category. However, it is smaller than the effects of PCs brought to the fore, determined by factors like the population median age, the epidemic onset, and the net immigration. Thereby, the diversity of the variables constituting PC1 in the supervised PCA, together with multiple GHSI categories, additionally questions the GHSI significance, as it makes it hard to separate the influence of the GHSI from the influences of other variables related to the country development level (HDI/GDP). Finally, the variable importance analysis points to a robust and dominant influence of the age PC1, while a positive influence of the GHSI Detect category is present but almost negligible compared to the age PC1 effect.

Moreover, the Random Forest method selects the Onset as an additional variable with higher significance than the GHSI Detect. Therefore, our results show that oversimplified analytical methods play a major role in the puzzle: While simple pairwise correlations indeed strongly suggest that “better healthcare leads to higher COVID-19 mortality”, there is hardly any hint of such paradox with advanced machine-learning analyses. Instead, old age and an earlier epidemic onset are likely behind the increased disease severity - and simple univariate analysis fails to identify these confounding variables, both significantly correlated with the GHSI.

Three severity predictors robustly selected by different methods – the median age, the epidemic onset, and the net immigrations – are generally supported by other studies’ results, obtained in combinations with other predictors ^6,8,11,13,14^. The fact that our analysis singled them out argues about their dominant roles. Old age is undoubtedly a dominant risk factor for developing severe illness and dying from COVID-19 ^6,8,13,14^. The onset variable, not acknowledged only by the Random Forest, is obtained with a negative association to severity ^11,37^. The time in which the pandemic reached a country may have been a significant factor in the first wave, as countries could use it to organize medical resources and adjust treatment protocols, learning from the experiences of those affected earlier. The Elastic net and the supervised PCA results identify the net immigration as a negative contributor to the COVID-19 severity. The supervised PCA groups this variable inside the same PC with the percent of refugees. This negative effect of immigration in the broader sense on the severity should not be interpreted through a hypothetical mitigating effect of the full lockdowns on the virus spread, rapidly imposed in those countries expecting a massive inflow of people ^13,38^, because m/r is independent of the disease transmissibility. Rather than being more efficient in treating COVID-19, countries leading in net immigration probably had an additional under-privileged population whose deaths from COVID-19 were more frequently misattributed to other causes, e.g., a lack of medical insurance or a general reluctance to go to hospitals ^39,40^. If so, the presence of immigrants did not decrease the actual COVID-19 mortality, but only the officially recognized toll (further discussed below).

We further demonstrate that the reliability of COVID-19 data also plays an important role. Many factors can lead to underreporting, or even overreporting, of COVID-19 deaths: testing capabilities, protocols for classifying causes of death, to outright political influences ^41^. Since these factors can be highly variable between countries, we considered the overall excess deaths as an alternative measure to official COVID-19 mortality statistics. We obtain that, similarly to ^42^, the excess deaths variable depends almost only on the sheer number of COVID-19 cases (itself not significantly correlated with any other of our predictor variables, see Fig. 1), together with some influence of the CFR value (where CFR itself is dominantly correlated with median age, and with epidemic onset - possibly due to the accumulated experience in medical treatments). The counterintuitive connection of COVID-19 severity with GHSI categories (and country’s prosperity) does not appear when considering the overall excess mortality. Another perspective on the phenomenon is gained by considering the “unexplained deaths” variable, quantifying the part of the overall excess deaths not officially attributed to COVID-19 mortality. We obtained that unexplained deaths are smaller in developed countries with high GHSI scores. Other studies also found a similar relationship between unaccounted excess deaths and the lower average income/fewer physicians per capita ^6,8,14,40,41,43^. This strongly suggests that underreporting COVID-19 deaths plays another crucial role in the GHSI puzzle, i.e., that the actual COVID-19 toll in low GHSI countries is higher than the official figures reveal.

For our present purposes, it is not crucial whether the excess deaths represent direct COVID-19 victims or indirect pandemic causalities of reduced access to healthcare services (i.e., due to overwhelmed medical facilities): GHSI scores should reflect capabilities to cope with both aspects of the pandemic consequences. Nevertheless, most of the unaccounted deaths likely represent direct consequences of SARS-CoV-2 infection, especially in countries where the difference between the official COVID-19 mortality and the excess of deaths is substantial. Namely, our results are based on the excess of deaths that occurred during a relatively short time window of the first pandemic wave (a few months), where the negligence of other/chronic medical conditions (e.g., postponed oncological or cardiological check-ups and treatments) will unlikely have such immediate effects on mortality. Besides, if the unexplained deaths were primarily an indirect consequence of saturation of medical resources, COVID-19 case counts should reflect this saturation and thus be expected to appear as a relevant predictor of unexplained deaths - but this does not happen. On similar grounds, the authors of ^42^ support the same interpretation of excess mortality in regions of Italy. As an additional argument, we note that, of all GHS indices, both the linear regressions with feature selection (Fig. 2) and the machine learning techniques (Fig. 4) consistently single out the Detect score (quantifying testing capabilities) as the one most affecting m/r. This is consistent with the interpretation that high GHSI scores - and the Detect index category in particular - are not connected to higher COVID-19 mortality but rather reflect better capacity to recognize (i.e., “detect”) COVID-19 deaths as such, and thus lead to less unexplained deaths. On the other hand, a lower Detect score relates to a larger number of unrecognized COVID-19 deaths and falsely lower mortality, adding to the apparent paradox.

We have provided ample arguments that the puzzling connection of high GHSI scores with severe COVID-19 epidemic outcomes is an artifact of oversimplified analyses and low-quality data, but neither of our methods could directly establish the anticipated negative correlation. Notably, the negative correlation with the scale of under-reporting of COVID-19 deaths is suggested by our results, demonstrating the effectiveness of better detection capacity of high-GHSI countries. However, it seems that the anticipated benefits of the elaborated preparedness capacities for minimizing the overall harm did not manifest, at least not directly. Note that our results clearly show the strong influence of population age and, in the second place, the epidemic onset on the m/r measure of severity. The lather predictor emphasizes the particular vulnerability of the initial pandemic period ^11,12^. Not only did a later onset provide more time to organize a response, but the large unpredictability effect when the world encountered an unknown virus, obstructing any organizational efforts ^1,44,45^, put the first-hit countries in an additionally vulnerable position. The lack of knowledge and experience could have primarily determined the reach of the initial efforts to minimize severity regardless of the pre-established capacities, and this factor would be hard to incorporate in a static preparedness measure such as the GHSI. Nevertheless, the GHSI might show a better, anticipated agreement later during the pandemic, when the advantages of the prepared system start to reveal. Previous studies, like ours, assessed the GHSI predictivity focusing on the pre-vaccination period, so its performance separated from the sensitive, first outbreak period remains largely unexplored.

On the other hand, GHS indices are positively correlated with the population age and negatively correlated with the onset time. In turn, if there was no direct influence of GHSI scores on mortality (nor additional confounding correlations), we should expect more excess deaths in countries with higher GHS indices - simply because the population is, on average, older, while the epidemic came earlier, in these countries. However, since the plot in Fig. 6 does not reveal such a trend, and overall excess mortality seems to depend almost solely on the number of infected individuals (with neither age nor GHSI playing any role in Fig. 5), there must be a factor that compensates for this age and early epidemic onset disadvantage. While we cannot rule out possible confounders, it is plausible that the difference in levels of medical care - reflected via GHSI scores - counterbalances these two effects.

## Conclusion

The main goal of this study was to reinvestigate the “GHSI puzzle,” giving attention to three essential aspects i) the choice of the effective epidemic burden measure, ii) the advanced multivariate data analysis, suiting the data complexity, and iii) the analysis of excess deaths data, in addition to more questionable official COVID-19 infection and mortality figures. Our results point to a major weak point in the previous efforts to assess the predictivity of the GHS index – namely, using the oversimplified methodology, which emphasizes the importance of implementing the three essential steps in our analysis mentioned above. We show that high GHSI scores cannot be associated with low efficacy in combating epidemics, contrary to the dominant claim in academic literature. On the contrary, more developed countries with high GHSI values likely provided better quality data due to more reliable reporting of COVID-19 fatality counts. This becomes particularly significant in light of the notion strongly supported by our results: that a majority of excess deaths after the first wave should be attributed to direct COVID-19 fatalities instead of indirect consequences of the pandemic or other concurrent causes.

GHSI categories, therefore, do not contradict the observed data and can still be a valuable instrument in ameliorating the consequences of future global health-related crises, especially with some refinement to account for the country’s vulnerability to a high level of initial unpredictability when it faces an unknown, fast-transmitting virus. Refining this and developing similar measures can aid the prevention of future epidemic catastrophes, where our study may provide some guidelines on how to perform an accurate assessment of their appropriateness.

## Data Availability

We confirm that all methods were carried out in accordance with relevant guidelines and regulations as publicly available data set is used.
All data generated or analyzed during this study are included in this article and its Supplementary material.

## Statements

We confirm that all methods were carried out in accordance with relevant guidelines and regulations as publicly available data set is used.

All data generated or analysed during this study are included in this published article and its Supplementary material.

