## Supplementary Material for "GHSI COVID-19 puzzle: did highly developed countries indeed fare worse?"

#### Data collection and processing

The majority of the data used in this research, including 18 demographic and health parameters, and the basic reproduction number of the virus was taken from the dataset previously assembled in (1). Additionally, Global Health Security Index (GHSI) data were taken from (2). In addition to the main (Overall) GHSI and six index categories (Prevent, Detect, Respond, Health, Norms, and Risk), the Covid index was calculated from the averaged values of COVID-19 related GHSI indicators, according to (3). COVID-19 counts (average and maximal daily cases and deaths) were taken from (4). At the end of the first peak, the case fatality rate and severity measure (m/r) were calculated according to the methodology described in (5). To determine the numbers of total and excess deaths during the first peak of the pandemic, data from (6) was used. For the research two datasets were constructed: *i)* m/r dataset - a subset of the dataset used in (1), consisting of the data from 85 countries for which both GHSI were available and m/r was inferred; *ii)* excess deaths dataset - a subset of *i)* for which total and excess deaths data was available (59 countries). Since the distribution of most variables initially deviated from normality, data was transformed, so that the skewness of the variables was as close to zero as possible, with the minimal number of remaining outliers. To obtain the best possible results, transformations were applied separately for variables in each dataset. Interpretation of all the variables and the used transformation for both datasets are presented in Supplement Table 1.

#### Principal Component Analysis

To partially decorrelate data and reduce dimensionality while retaining relatively simple interpretation of principal components, some of the variables were grouped into mutually related groups, on which Principal Component Analysis (PCA) was performed. The number of principal components retained from each group after the PCA was determined to explain over 85% of the variance of the data. PCA grouping was performed independently for both datasets and variables entering PCA, retained principal components, and the percentage of variance they account for are presented in Supplement table 2 (m/r dataset) and 3 (excess deaths dataset). New datasets, now consisting of both retained principal components and remaining variables that did not enter PCA, were next used as input in univariate and multivariate analyses. For easier interpretation of the most relevant principal components, their correlation with the variables entering PCA is given in Supplement Figures 1 and 4. Two now obtained sets of predictors were: i) variables used in m/r regression analysis, consisting of: Detect, Respond, and Health GHSI categories, five selected PCs from Supplement table 2, and remaining demographic and health parameters that did not enter PCA (BUAPC, UP, IE, RE, OB, SM, IN, BCG, ON) ii) variables used in excess and unexplained deaths analysis with principal components consisting of $R_0$, CFR and 11 principal components from Supplementary Table 3.

#### Linear regression models

LASSO and Elastic net regressions were used as the implementations of L1 (LASSO) and L1 and L2 norms (Elastic net) on three variations of the m/r dataset and on excess deaths dataset with principal components (regressions were implemented separately for excess and unexplained deaths as response variables). The variables (and PCs) were standardized before the regressions, so the regression coefficients obtained by these methods can be interpreted as the relative importance of predictors in explaining the response variables. Values of the hyperparameters $\lambda$ in LASSO and $\alpha$ and $\lambda$ in Elastic Net were determined through 5-fold cross-validation, with the data repartitioned 40 times. In m/r dataset regressions, the sparsest model - with mean squared error (MSE) within 1 standard error (SE) from the minimal MSE - model was chosen. To further reduce the noise in m/r dataset regressions (where the number of predictors retained after the first round of regression was relatively high), LASSO and Elastic Net were implemented as "Relaxed" (7), meaning that the regression was performed in two rounds so that only the variables selected in the first round were used as the input for the second round of regression. Regressions were performed on the initial m/r dataset, without grouping and PCA and with Detect, Respond, Health and Risk GHSI categories (Supplement Figure 2); on a dataset containing Covid index and 18 demographic and health variables, without any other GHSI categories (Supplement Figure 3); and on m/r dataset with principal components (Figure 2). Since the number of selected variables after the first round of regression with both excess and unexplained deaths data was relatively small (even though this time minimal MSE model was selected instead of the sparsest), the second round of regression was not performed. As an addition to LASSO and Elastic Net for the excess and unexplained deaths analysis, Forward stepwise linear regression (8) was performed. In this method, starting from the constant model, a predictor is added to the regression if it improves the fit significantly ($P < 0.05$ in F-statistics). The process is repeated sequentially, adding significant and removing non-significant terms until the best fit is obtained.

### Random Forest and Gradient Boost regressions

Two non-parametric, decision tree-based methods – Random Forest (8,9) and Gradient Boost (8,10) - were implemented on both m/r and excess deaths datasets with principal components. These methods are of particular importance, as they can accommodate potential interaction between the variables and highly non-linear relations between the predictors and the response variable. In the analysis of disease severity, the m/r dataset with principal components was used, with a constraint that only variables correlated to m/r with P<0.1 in either Pierson, Kendall, or Spearman correlation are used as the input in the model. This way, overfitting, which would present a problem for these algorithms, is avoided. As the number of input variables was smaller in the excess/unexplained deaths dataset, preselection was not required before those regressions. Values of the hyperparameters in Random Forest and Gradient Boost methods were selected through extensive grid search, by the same method of cross-validation used for LASSO and Elastic Net regressions, and a model with the minimal MSE was chosen to be retained on the entire (reduced) dataset.

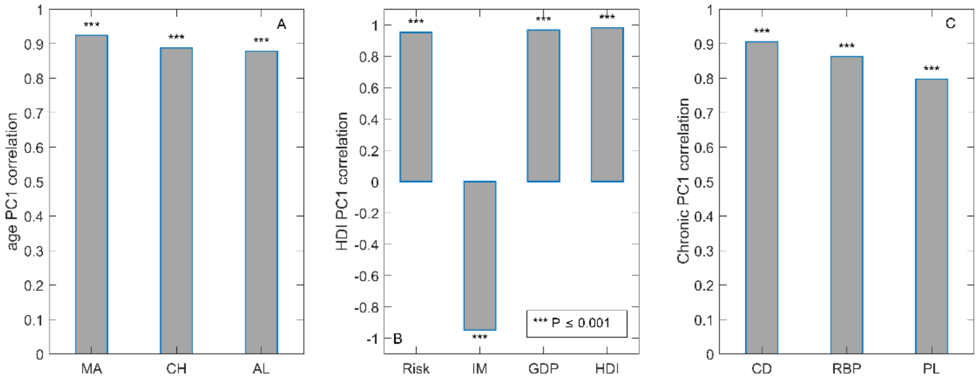


**Supplement Fig. 1: Correlations of relevant principal components with variables entering PCA for m/r dataset**. MA – median age, CH – blood cholesterol level, AL – alcohol consumption, Risk – GHSI Risk category, IM – infant mortality, GDP – gross domestic product per capita, HDI – human development index, CD – prevalence and severity of chronic diseases, RBP – prevalence of raised blood pressure, PL – long-term $PM_{2.5}$ pollution.

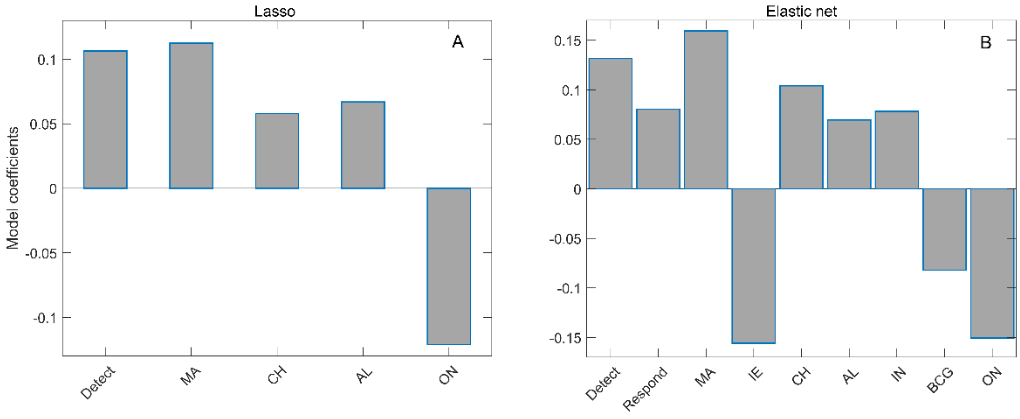


**Supplement Fig. 2: Relaxed Lasso (A) and Relaxed elastic net (B) regressions on the initial set of demographic variables and Detect, Respond, Health and Risk GHSI categories**. Detect – GHSI Detect category, Respond – GHSI Respond category, MA – median age, IE – net immigration, CH – blood cholesterol level, AL – alcohol consumption, IN – prevalence of physical inactivity, BCG – BCG immunization coverage, ON – epidemic onset.

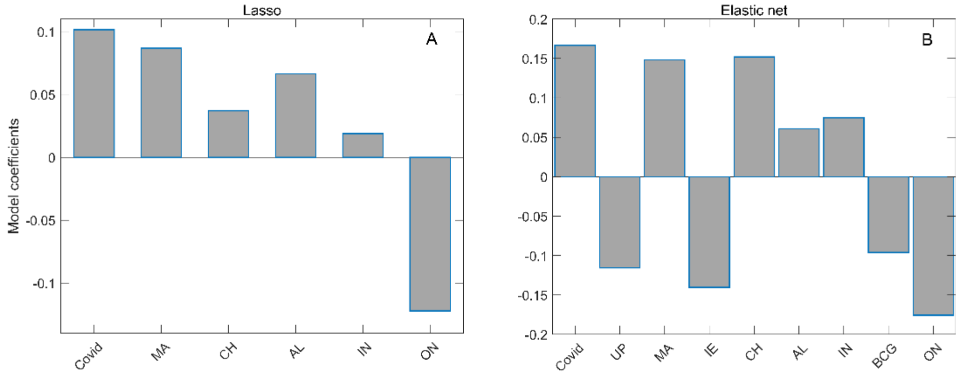


**Supplement Fig. 3:** LASSO (A) and Elastic net (B) on the initial set of demographic variables and Covid index. Covid – GHSI Covid category, UP – urban population, MA – median age, IE – net immigration, CH – blood cholesterol level, AL – alcohol consumption, IN – prevalence of physical inactivity, BCG – BCG immunization coverage, ON – epidemic onset.

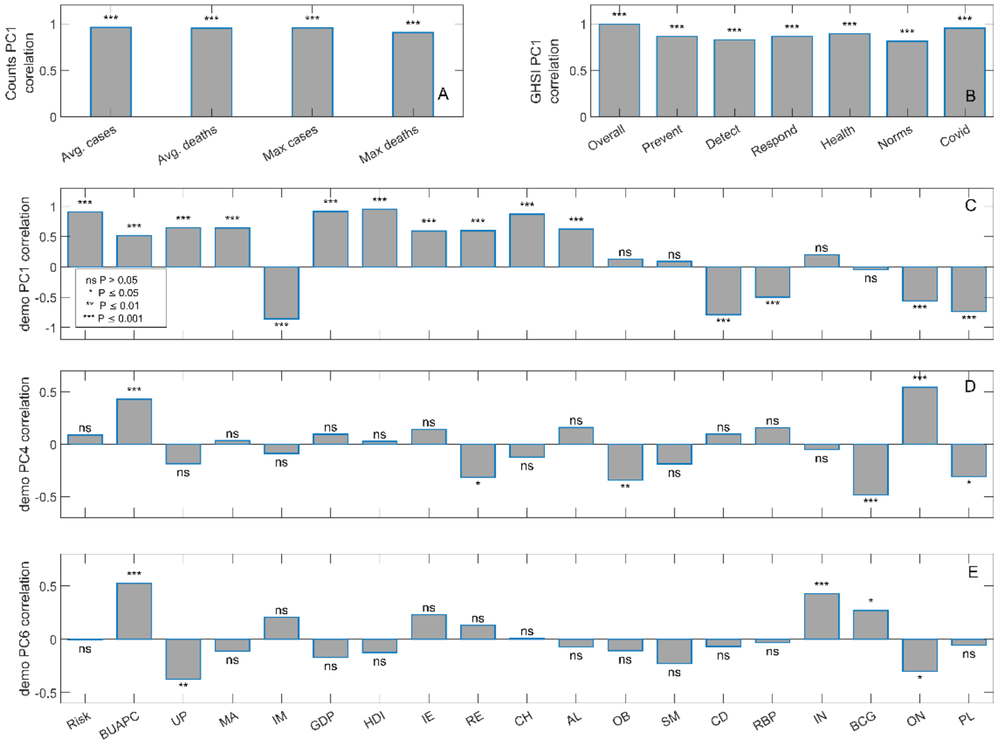


**Supplement Fig. 4: Correlation of relevant principal components with the variables entering PCA in excess deaths dataset**. Variable interpretation: A) Avg. cases- average daily COVID-19 cases, Avg. deaths - average daily COVID-19 deaths, Max cases- maximal daily number of cases, Max deaths – maximal daily COVID-19 deaths B) GHSI indices C-E) Risk – GHSI Risk index, BUAPC – built-up area per capita, UP – urban population, MA – median age, IM – infant mortality, GDP – gross domestic product per capita, HDI – human development index, IE – net migration, RE – refugees, CH – blood cholesterol level, AL – alcohol consumption, OB – prevalence of obesity, SM – prevalence of smoking, CD – prevalence of cardiovascular diseases, RBP – raised blood pressure, IN – physical inactivity, BCG – BCG vaccination coverage, ON – the onset of the epidemic, PL – air pollution.

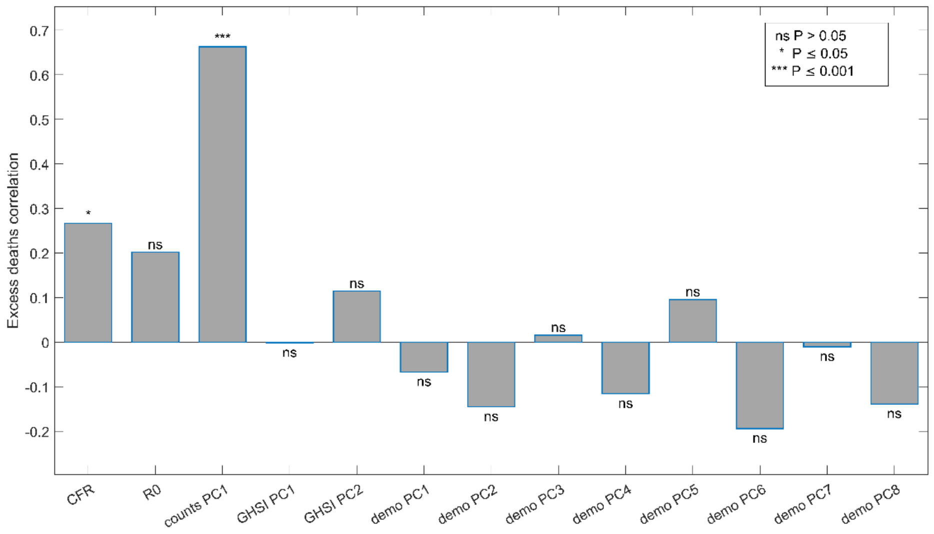


**Supplement Fig. 5:** Pearson's Correlation of excess deaths and selected variables and principal components. CFR – case fatality rate, R0 – basic reproduction number of the virus. For principal components explanation, see Supplementary Table 3.

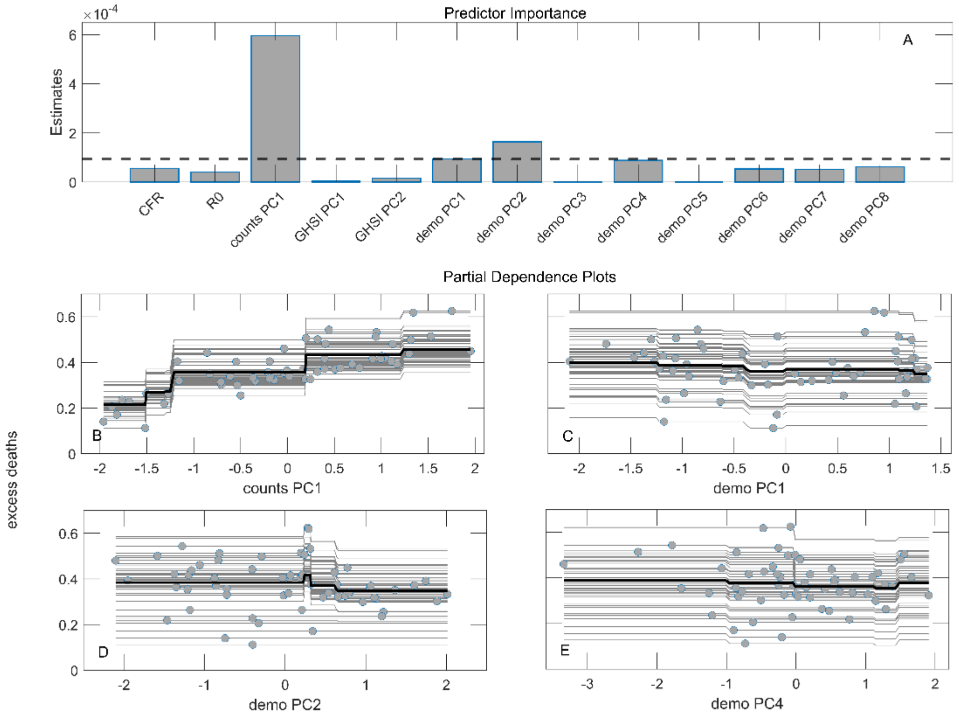


**Supplement Fig. 6:** Gradient Boost regression with excess deaths as the response variable. A) Predictor importance estimates. The dashed line represents the mean value of predictor importance. CFR – case fatality rate, R0 – basic reproduction number of the virus. For principal components explanation, see Supplementary Table 3. B-E) Partial dependency plots for selected variables with the highest importance estimates.

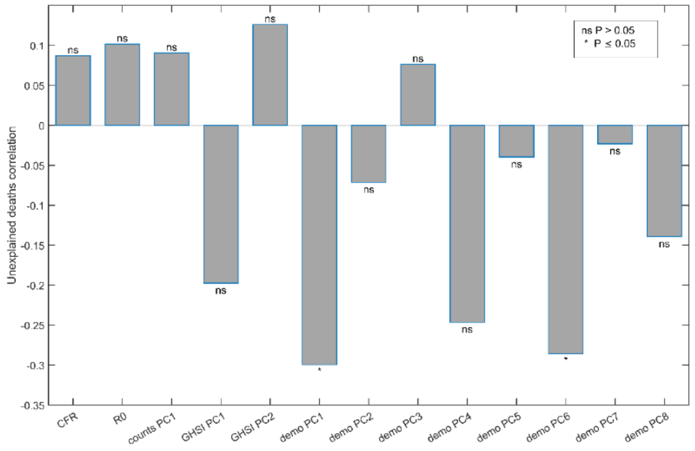


**Supplement Fig. 7:** Pearson's Correlation of unexplained deaths and selected variables and principal components. CFR – case fatality rate, R0 – basic reproduction number of the virus. For principal components explanation, see Supplementary Table 3.

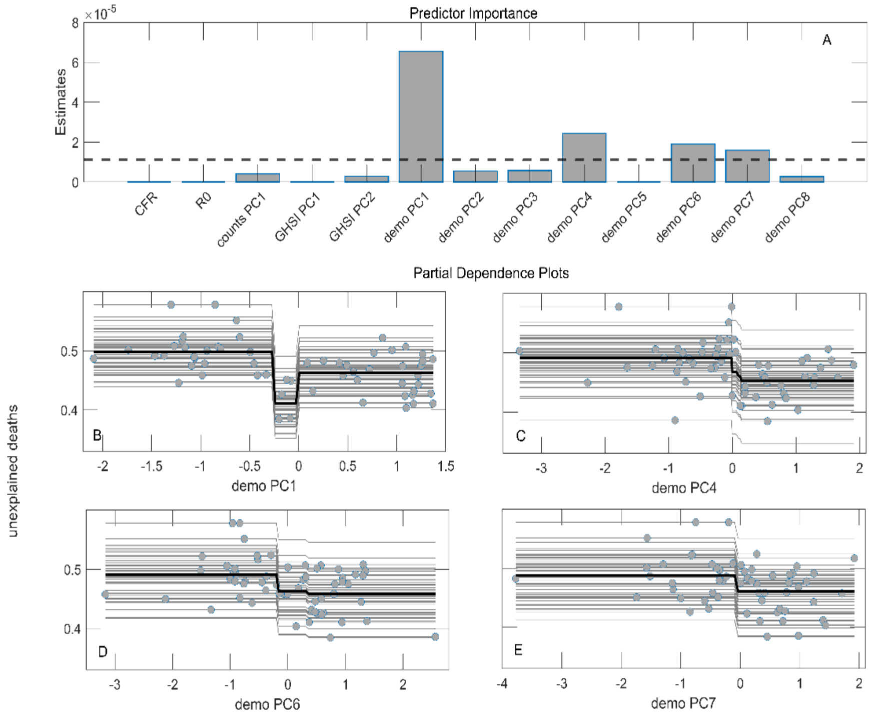


**Supplement Fig. 8:** Gradient Boost regression with unexplained deaths as the response variable. A) Predictor importance estimates. The dashed line represents the mean value of predictor importance. CFR – case fatality rate, R0 – basic reproduction number of the virus. For principal components explanation, see Supplementary Table 3. B-E) Partial dependency plots for selected variables with the highest importance estimates.

#### Supplement Table 1: Data transformation

| Variable | Abbreviation | Transformation in m/r dataset | Transformation in excess deaths dataset |
|---|---|---|---|
| Disease severity measure | m/r | $\log(x)$ | / |
| Relative excess deaths | Excess deaths | / | $\sqrt{x-\min(x)}$ |
| Relative unexplained deaths | Unexplained deaths | / | $\sqrt[3]{x-\min(x)}$ |
| Case fatality rate | CFR | / | $\log(x)$ |
| Basic reproduction number of SARS-CoV-2 | R0 | / | $\log(x)$ |
| Average daily cases | Avg. cases | / | $\log(x)$ |
| Average daily deaths | Avg. deaths | / | $\log(x)$ |
| Max daily cases (per capita) | Max cases | / | $\log(x)$ |
| Max daily cases (per capita) | Max deaths | / | $\log(x)$ |
| Overall GHSI | Overall | $\sqrt[3]{x}$ | $\sqrt[3]{x}$ |
| Prevent GHSI category | Prevent | None | None |
| Detect GHSI category | Detect | None | None |
| Respond GHSI category | Respond | $\sqrt[3]{x}$ | $\sqrt[3]{x}$ |
| Health GHSI category | Health | None | None |
| Norms GHSI category | Norms | None | None |
| Risk GHSI category | Risk | $x^2$ | None |
| COVID-19 index | Covid | $\sqrt{x}$ | $\sqrt{x}$ |
| Built-up area per capita | BUAPC | $\sqrt{x}$ | $\log(x)$ |
| Urban population | UP | $x^2$ | $x^2$ |
| Median age | MA | None | $-\log(\max(x)-x)$ |
| Infant mortality | IM | $\log(x)$ | $\log(x)$ |
| Gross domestic product per capita | GDP | $\log(x)$ | $\log(x)$ |
| Human development index | HDI | $-\sqrt{\max(x)-x}$ | $x^2$ |
| Net immigration | IE | $-\sqrt{\max(x)-x}$ | None |
| Percentage of refugees | RE | $\log(x)$ | $\log(x)$ |
| Average blood cholesterol level | CH | $-\sqrt{\max(x)-x}$ | $x^2$ |
| Alcohol consumption | AL | None | $x^2$ |
| Prevalence of obesity | OB | $-\log(\max(x)-x)$ | $\sqrt[3]{x}$ |
| Prevalence of smoking | SM | None | None |
| Prevalence and severity of chronic diseases | CD | $\sqrt[3]{x}$ | $\log(x)$ |
| Prevalence of raised blood pressure | RBP | $\sqrt{x}$ | $\log(x)$ |
| Prevalence of insufficient physical activity | IN | $-\sqrt{\max(x)-x}$ | None |
| BCG immunization coverage | BCG | $-\sqrt{\max(x)-x}$ | $-\sqrt{\max(x)-x}$ |
| Epidemic onset (days from 15.02.2020. until the beginning of the epidemic in the given country) | ON | $\log(x)$ | None |
| Long-term average $PM_{2.5}$ pollution | PL | $\log(x)$ | $\log(x)$ |

“ / ” – indicates that variable was not included in the dataset, “None” – no transformation was applied.

#### Supplement Table 2: grouping of variables and PCA for m/r dataset

| Variables entering PCA | Retained principal components | Variance explained |
|---|---|---|
| Risk | HDI PC1 | 92.7% |
| Infant Mortality | | |
| GDP per capita | | |
| Human development index | | |
| Median age | Age PC1<br>Age PC2 | 92.5% |
| Blood cholesterol level | | |
| Alcohol consumption | | |
| Chronic diseases | Chr PC1<br>Chr PC2 | 91.0% |
| Raised blood pressure | | |
| Pollution | | |

HDI PC1 – Human development index principal component 1, Chr PC1,2 – Chronic diseases principal components

#### Supplement Table 3: grouping of variables and PCA for excess deaths dataset

| Variables entering PCA | Retained principal components | Variance explained |
|---|---|---|
| Average Daily Cases | Counts PC1 | 89.0% |
| Average Daily deaths | | |
| Max daily cases (per capita) | | |
| Max daily cases (per capita) | | |
| Overall GHSI | GHSI PC1<br>GHSI PC2 | 85.6% |
| Prevent | | |
| Detect | | |
| Respond | | |
| Health | | |
| Norms | | |

| | | |
|---|---|---|
| Covid | | |
| Built-up area per capita | Demo PC1<br>Demo PC2<br>Demo PC3<br>Demo PC4<br>Demo PC5<br>Demo PC6<br>Demo PC7<br>Demo PC8 | 86.8% |
| Urban population | | |
| Median age | | |
| Infant mortality | | |
| GDP per capita | | |
| Human development index | | |
| Immigrants – Emigrants | | |
| Refugees | | |
| Blood cholesterol level | | |
| Alcohol Consumption | | |
| Obesity | | |
| Smoking | | |
| Chronic diseases | | |
| Raised blood pressure | | |
| Physical inactivity | | |
| BCG vaccination coverage | | |
| Epidemic onset | | |
| Air pollution | | |
| Risk | | |

### Supplement Table 4: Excess deaths linear regression models

**Stepwise linear regression**

| Predictor | Estimate | SE | tStat | pValue |
|---|---|---|---|---|
| Counts PC1 | 0.10 | 0.015 | 6.7 | $10^{-8}$ |

$R^2 = 0.44$, Adjusted $R^2 = 0.43$
P-value = $10^{-8}$

**Lasso regression**

| Predictor | Estimate |
|---|---|
| Counts PC1 | 0.072 |

$\lambda = 0.03$
min MSE = 0.74, SE min MSE = 0.032
$R^2 = 0.40$

**Elastic Net regression**

| Predictor | Estimate |
|---|---|
| Counts PC1 | 0.073 |

$\alpha = 0.73$
$\lambda = 0.04$
min MSE = 0.72, SE min MSE = 0.036
$R^2 = 0.41$

### Supplement Table 5: Unexplained deaths linear regression models

**Stepwise linear regression**

| Predictor | Estimate | SE | tStat | pValue |
|---|---|---|---|---|
| R0 | 0.028 | 0.013 | 2.2 | 0.03 |
| Demo PC1 | -0.039 | 0.012 | -3.1 | 0.003 |
| Demo PC6 | -0.033 | 0.012 | -2.8 | 0.007 |

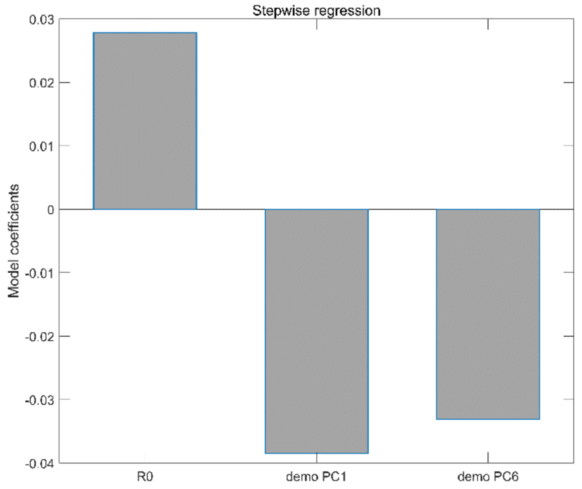


$R^2 = 0.24$, Adjusted $R^2 = 0.20$
P-value = 0.002

**Lasso regression**

| **Predictor** | **Estimate** |
|---|---|
| Demo PC1 | -0.009 |
| Demo PC4 | -0.004 |
| Demo PC6 | -0.008 |

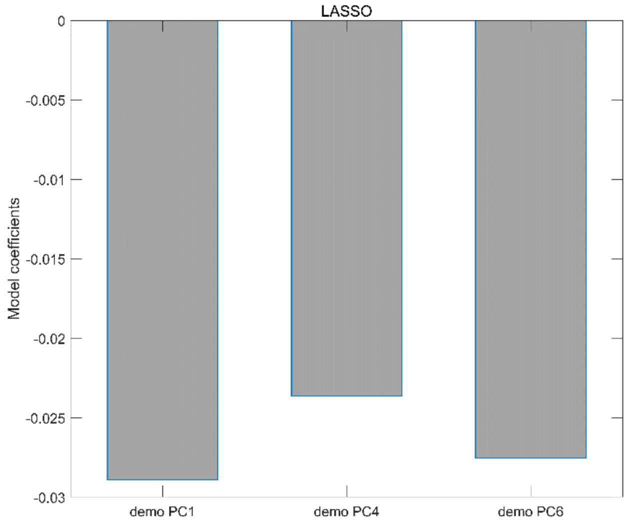


$\lambda = 0.02$
min MSE = 1.1 , SE min MSE = 0.030
$R^2 = 0.10$

**Elastic net regression**

| **Predictor** | **Estimate** |
|---|---|
| Demo PC1 | -0.011 |
| Demo PC4 | -0.0058 |
| Demo PC6 | -0.0097 |

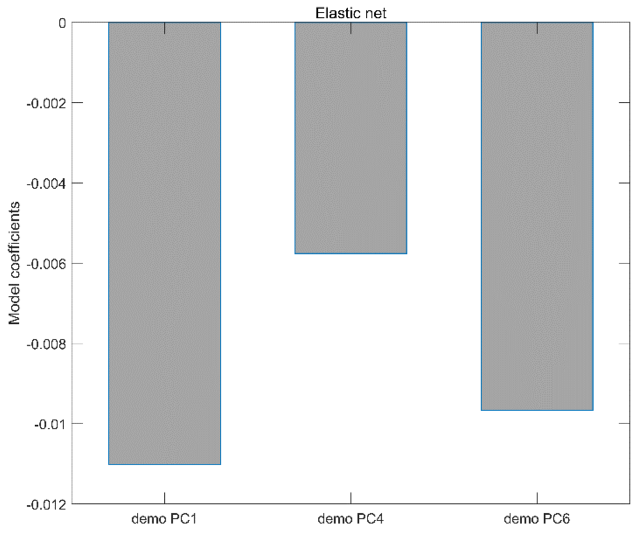


α = 0.83
λ = 0.02
min MSE = 1.12, SE min MSE = 0.030
$R^2$ = 0.13
